# Evaluating the Clinical Feasibility of an Artificial Intelligence-Powered Clinical Decision Support System: A Longitudinal Feasibility Study

**DOI:** 10.1101/2021.07.03.21259812

**Authors:** Christina Popescu, Grace Golden, David Benrimoh, Myriam Tanguay-Sela, Dominique Slowey, Eryn Lundrigan, Jérôme Williams, Bennet Desormeau, Divyesh Kardani, Tamara Perez, Colleen Rollins, Sonia Israel, Kelly Perlman, Caitrin Armstrong, Jacob Baxter, Kate Whitmore, Marie-Jeanne Fradette, Kaelan Felcarek-Hope, Ghassen Soufi, Robert Fratila, Joseph Mehltretter, Karl Looper, Warren Steiner, Soham Rej, Jordan F. Karp, Katherine Heller, Sagar V. Parikh, Rebecca McGuire-Snieckus, Manuela Ferrari, Howard Margolese, Gustavo Turecki

## Abstract

**Objective:** We examine the feasibility of an Artificial Intelligence (AI)-powered clinical decision support system (CDSS), which combines the operationalized 2016 Canadian Network for Mood and Anxiety Treatments guidelines with a neural-network based individualized treatment remission prediction.

**Methods:** Due to COVID-19, the study was adapted to be completed entirely at a distance. Seven physicians recruited outpatients diagnosed with major depressive disorder (MDD) as per DSM-V criteria. Patients completed a minimum of one visit without the CDSS (baseline) and two subsequent visits where the CDSS was used by the physician (visit 1 and 2). The primary outcome of interest was change in session length after CDSS introduction, as a proxy for feasibility. Feasibility and acceptability data were collected through self-report questionnaires and semi-structured interviews.

**Results:** Seventeen patients enrolled in the study; 14 completed. There was no significant difference between appointment length between visits (introduction of the tool did not increase session length). 92.31% of patients and 71.43% of physicians felt that the tool was easy to use. 61.54% of the patients and 71.43% of the physicians rated that they trusted the CDSS. 46.15% of patients felt that the patient-clinician relationship significantly or somewhat improved, while the other 53.85% felt that it did not change.

**Conclusions:** Our results confirm the primary hypothesis that the integration of the tool does not increase appointment length. Findings suggest the CDSS is easy to use and may have some positive effects on the patient-physician relationship. The CDSS is feasible and ready for effectiveness studies.

## Introduction

Clinical decision support systems (CDSS) consolidate large quantities of clinical information in order to provide clinicians with the necessary data to support medical decision-making and assist with managing treatment protocols (Zikos & Delellis, 2018; Sutton et al., 2020). One emerging focus of medical informatics is the improvement of patient care through data-driven, patient-centred decision support systems. Artificial intelligence (AI) algorithms are increasingly being integrated into CDSS, permitting predictive analytics to be used by clinicians as part of routine practice (Sutton et al., 2020). The overarching objective of these systems is the improvement of medical decision-making using a data-driven approach. However, while much has been written about the machine learning techniques (Meltretter et al., 2019; Squarcina et al., 2021) which underpin the technical advancements that make these systems possible, comparatively less focus has been placed on the useability and feasibility of these kinds of systems in medicine in general, and in mental health treatment in particular. In this paper, we will discuss a feasibility study of a novel AI-powered CDSS aimed at improving depression treatment.

Feasibility and ease of use is a major concern as it roughly equates to the tolerability of a drug treatment, with similar impact-much like a medication, a digital tool can only have a positive impact if patients (and, in this case, clinicians) use it and continue to use it. A recent meta-analysis of randomized controlled trials aimed to establish the dropout rates of studies of medical smartphone apps tracking depressive symptoms (Torous et al., 2020). The analysis found that apps for depressive symptom tracking had a dropout rate of approximately 50% when accounting for bias. Despite this high dropout rate, there is some knowledge about how to reduce dropout. For example, researchers found that the dropout rate was significantly lower in apps offering human feedback and in-app mood monitoring -as low as 12% (Torous et al., 2020). Additionally, previous decision support systems have demonstrated a need for a tool that provides real-time utility (Rollman et al., 2002), the ability to personalise treatment choices and differentiate between medications (Harrison et al., 2020) in a quantifiable manner (Adli et al., 2017), and incorporate clinical practice guidelines (Benrimoh et al., 2020).

Only one-third of depression patients who receive treatment will achieve remission during the first treatment course; the majority experience multiple treatment trials before entering remission (Warden et al., 2007). Clinicians are faced with a wide range of treatment options, in combination with associated guidelines, to manage selected treatments. However, there are no easily accessible, point-of-care tools available to aid in the optimization of treatment success and minimize time to remission. Furthermore, treatments are essentially equally effective at the population level, when to improve outcomes treatment selection must address the individual’s specific characteristics (Cipriani et al., 2018; Benrimoh et al., 2020). As such, there is a clear need for improved and personalized decision support for mental healthcare (see Benrimoh et al., 2018 for further discussion).

Aifred is a clinical decision support system (CDSS) that uses artificial intelligence (AI) to assist clinicians in selecting treatments for MDD. The tool incorporates a deep-learning model, validated and trained on clinical and demographic baseline data in order to support treatment selection by providing individualized probabilities of remission for specific treatment options (see Benrimoh et al., 2020, Mehltretter et al., 2020a & 2020b for a description of the tool and machine learning model, and model training and validation methodology). The tool integrates an operationalized version of the 2016 Canadian Network for Mood and Anxiety Treatments (CANMAT) guidelines for depression treatment (Kennedy et. al., 2016) and allows physicians to track their patient’s symptoms using standardized questionnaires and to visualize this progress using graphed data, supporting the implementation of measurement based care and algorithm guided treatment. The AI aspect of the CDSS is directly integrated into the operationalized CANMAT guidelines, with the remission probabilities for individual treatments presented within the guideline module when antidepressant treatments are being chosen. The application is designed to support clinicians by considering complex interactions between multiple patient clinical, social, and demographic variables, to help personalize treatment in order to improve upon a trial-and-error treatment approach and reduce the number of failed treatment trials (Benrimoh et al., 2020). To summarize, the application assists clinicians in providing measurement-based, treatment-algorithm guided and AI-personalized care.

Patients also have access to their own version of the application wherein they respond to questionnaires and can view their active and past treatments and their symptoms graphed over time. This availability of data to both physician and patient is intended to empower patients, enrich conversations, and facilitate shared decision making (Benrimoh et al., 2020).

Following a previous simulation center study (Benrimoh et al., 2020), and ahead of larger clinical trials aimed at assessing safety and effectiveness, we decided to conduct a feasibility study aimed at exploring the feasibility of the CDSS in a real clinical setting and to assess its longitudinal impact on the patient-clinician relationship. One key metric brought up by clinicians interviewed during initial stakeholder conversations was appointment length-clinicians are increasingly required to interact with time-consuming digital systems, and as such the fear of yet another system adding time to assessments is a reasonable one (Ventola, 2014). As such, we measured appointment length as a key numerical proxy to real-world feasibility.

### Aims of the Study

1. To assess the feasibility of the CDSS for use in clinical practice.
2. To assess physician and patient trust in the CDSS.
3. To assess the useability of the CDSS and study software and ensure that major limitations are identified and rectified prior to clinical trials.
4. To assess engagement with the application.

## Methods

The study was approved by the Research Ethics Boards of the Douglas Mental Health University Institute (Identifier: NCT04061642). All participants provided written informed consent to participate. The study was conducted according to ethical principles stated in the Tri-Council Policy Statement on the Ethical Conduct for Research Involving Humans (Tri-Council Policy Statement, 2005).

This was a single arm, naturalistic follow-up study aimed at assessing software usability and acceptability conducted between January and November 2020. It was not designed to assess the clinical effectiveness of the tool, which will be the focus of an upcoming clinical trial. It is important to note that physicians were provided access to the tool but were free to use the tool and its AI predictions or ignore it.

Several hypotheses were pre-specified prior to the study start. First, we hypothesized that there would not be a significant difference in measured appointment lengths between a baseline visit where the tool was not used and two post-baseline visits where the tool was used. Our second, related, hypothesis was that physicians would not subjectively report that using the CDSS and study software increased the length of their appointments. Our third hypothesis was that at least 66% of patients and 66% of physicians would rate the trustworthiness of the CDSS as a 4 or 5 on a 5-point Likert scale (with higher ratings indicating greater trust). Our fourth hypothesis was that at least 66% of patients and 66% of physicians would rate the overall usability of the CDSS as a 4 or 5 on a 5 point Likert scale (with higher ratings indicating greater usability). Our fifth hypothesis was that at least 70% of physicians and 65% of patients would still be using the application regularly by the end of the study. For physicians, regularly was defined as the application being used in every study-related visit. This was measured by checking if physicians logged into the application at each appointment. For patients, regularly was defined as completing at least one full PHQ-9 and GAD-7 questionnaire on the application per week.

The study sample included two population groups: 1) physicians, including family physicians and psychiatrists; and 2) patients of these physicians. The recruitment target was ten physicians and three to four patients per physician (30 to 40 patients total).

Physicians were recruited via a recruitment email and direct contact by study personnel. Sites consisted of university hospitals, primary care clinics and psychiatric clinics in the Canadian province of Québec. Eligible physicians were family physicians or psychiatrists treating patients with depression on at least a monthly basis. Physicians who met eligibility criteria were then invited to attend an introductory session with study personnel where the study and the AI model were described and training on how to use the tool was administered.

Participating physicians informed their patients with major depressive disorder (MDD) about the study and referred interested patients to study personnel. Eligible patients were patients of enrolled physicians who were at least 18 years of age and diagnosed with MDD by the physician as per DSM-V criteria (American Psychiatric Association, 2013), able and willing to provide informed consent, and not suffering from DSM-V diagnosed (or suspected) Bipolar Affective Disorder. Informed consent was obtained from patients and their accounts created and linked to that of their physician.

## Procedure

Upon account creation, patients were asked to complete the following questionnaires on the tool: Patient Health Questionnaire-9 (PHQ-9) to screen and track for depressive symptoms and their severity over time (Kroenke et al. 2001); General Anxiety Disorder-7 (GAD-7) to screen and track for anxiety symptoms and their severity over time (Spitzer et al. 2006); Alcohol Use Disorders Identification Test (AUDIT) to screen for harmful alcohol use (Bohn et al. 1995); Drug Abuse Screen Test (DAST-10) to screen for the presence and severity of problematic drug use; and Standardized Assessment of Personality -Abbreviated Scale Self Assessment (SAPAS-SA) to screen for personality disorders using a threshold of 3 points. The results of patient baseline questionnaire scores are summarized in Table 1, with clinical characteristics reported in Table 2.

**Table 1.** Patient clinical baseline scores.

| Questionnaire Baseline Score | Response Options | Frequency | Mean | Standard Deviation |
| --- | --- | --- | --- | --- |
| **SAPAS (n=15)**<br>This is a screening questionnaire for personality disorders, which helps your doctor understand if you may be suffering from a personality disorder or require further assessment. Interpreting the score | < 3 points: negative screen for personality disorder<br>≥ 3 points: positive screen for personality disorder. Further clinical evaluation is warranted. | (n=5)<br>33.33%<br>(n=10)<br>66.67% | 3.53 | 2.23 |
| **GAD-7 (n=14)**<br>This questionnaire helps screen for and track anxiety symptoms over time. Interpreting the score | 0-5: no or minimal anxiety<br>6-10: mild anxiety<br>11-15: moderate anxiety<br>16-21: severe anxiety | (n=2)<br>14.29%<br>(n=4)<br>28.57%<br>(n=2)<br>14.29%<br>(n=6)<br>42.86% | 12.21 | 5.81 |
| **PHQ-9 (n=15)**<br>This questionnaire helps screen for and track depression symptoms over time. Interpreting the score | 0-4: minimal or no depression<br>5-9: mild depression<br>10-14: moderate depression<br>15-19: moderately severe depression<br>20-27: severe depression | (n=1)<br>6.67%<br>(n=1)<br>6.67%<br>(n=4)<br>26.67%<br>(n=7)<br>46.67%<br>(n=2)<br>13.33% | 14.80 | 5.61 |
| **Audit (n=15)**<br>This is a screening questionnaire for problematic alcohol use, which helps your doctor understand if you might need further assessment or treatment for alcohol use. Interpreting the score | < 8: negative screen for harmful alcohol use<br>≥ 8: positive screen for harmful alcohol use | (n=12)<br>80.00%<br>(n=3)<br>20.00% | 4.40 | 3.58 |
| **DAST-10** (n=15)<br>This is a screening questionnaire for problematic drug use, which helps your doctor understand if you might need further assessment or treatment for drug use. Interpreting the score | 0: no problems reported<br>1-2: low level<br>3-5: moderate level<br>6-8: substantial level<br>9-10: severe level | (n=0)<br>0.00%<br>(n=11)<br>73.33%<br>(n=0)<br>0.00%<br>(n=3)<br>20.00%<br>(n=1)<br>6.67% | 2.73 | 2.89 |
| **WHODAS** (n=13)<br>The total score for this questionnaire ranges from 0-144. Higher scores indicate a lower ability to function. | n/a | n/a | 54.62 | 27.08 |
| **QIDS** (n=12)<br>A 16-item measure of depressive symptom severity | 1-5: no depression<br>6-10: mild depression<br>11-15: moderate depression<br>16-20: severe depression<br>21-27: very severe depression | (n=2)<br>16.67%<br>(n=2)<br>16.67%<br>(n=3)<br>25.00%<br>(n=4)<br>33.33%<br>(n=1)<br>8.33% | 13.12 | 6.04 |
| **IDS-C** (n=13) | 0-11: no depression<br>12-23: mild depression<br>24-36: moderate depression<br>37-46: severe depression<br>47-84: very severe depression | (n=0)<br>0.00%<br>(n=4)<br>30.77%<br>(n=5)<br>38.46%<br>(n=3)<br>23.08%<br>(n=1)<br>7.69% | 30.00 | 12.88 |

**Table 2.**
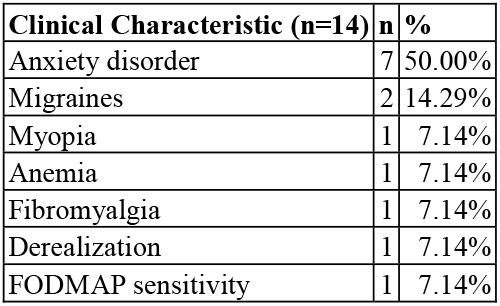
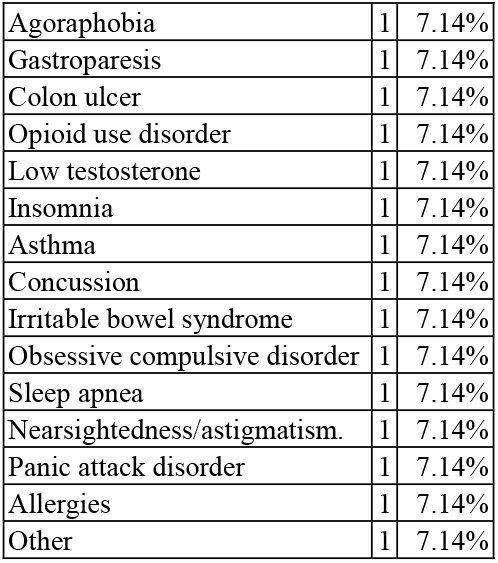
Patient clinical characteristics.

| **Clinical Characteristic (n=14)** | **n** | **%** |
| --- | --- | --- |
| Anxiety disorder | 7 | 50.00% |
| Migraines | 2 | 14.29% |
| Myopia | 1 | 7.14% |
| Anemia | 1 | 7.14% |
| Fibromyalgia | 1 | 7.14% |
| Derealization | 1 | 7.14% |
| FODMAP sensitivity | 1 | 7.14% |
| Agoraphobia | 1 | 7.14% |
| Gastroparesis | 1 | 7.14% |
| Colon ulcer | 1 | 7.14% |
| Opioid use disorder | 1 | 7.14% |
| Low testosterone | 1 | 7.14% |
| Insomnia | 1 | 7.14% |
| Asthma | 1 | 7.14% |
| Concussion | 1 | 7.14% |
| Irritable bowel syndrome | 1 | 7.14% |
| Obsessive compulsive disorder | 1 | 7.14% |
| Sleep apnea | 1 | 7.14% |
| Nearsightedness/astigmatism. | 1 | 7.14% |
| Panic attack disorder | 1 | 7.14% |
| Allergies | 1 | 7.14% |
| Other | 1 | 7.14% |

Patients were notified weekly by an automated email sent by the application to complete a PHQ-9, GAD-7, Patient Rated Inventory of Side Effects (PRISE-20) (to screen and track for specific antidepressant side effects and their severity), and Frequency, Intensity, Burden of Side Effects Rating (FIBSER) (to assess the overall impact of antidepressant side effects) (Wisniewski et al. 2006).

Between obtaining informed consent and their next visit with their physician, patients met with study personnel to complete a demographics questionnaire (Table 3), Adverse Childhood Experiences (ACE) questionnaire, and Life Events Checklist for DSM-5 (LEC-5) to screen for childhood or lifetime trauma (Felitti et al., 1998; Weathers et at., 2013).

**Table 3.** Patient demographics. 14 patients completed the study; however, 1 patient did not complete the demographic questionnaire.

| Patient Characteristics | Response options | N | (%) | Mean | Standard Deviation |
| --- | --- | --- | --- | --- | --- |
| **Age (years; n=14)** | N/A | 14 | N/A | 36.43 | 14.84 |
| **Gender (n=14)** | Male | 5 | 38.46% |  |  |
|  | Female | 9 | 69.23% |  |  |
| **Sex** | Male | 5 | 38.46% |  |  |
|  | Female | 9 | 69.23% |  |  |
| **Ethnicity (n=13)** | Caucasian | 10 | 76.92% |  |  |
|  | Caribbean | 1 | 7.69% |  |  |
|  | African or African American | 1 | 7.69% |  |  |
|  | Unanswered | 1 | 7.69% |  |  |
| **Adopted (n=13)** | Not adopted | 12 | 92.31% |  |  |
|  | Adopted | 1 | 7.69% |  |  |
| **Residency Status (n=13)** | Canadian citizen | 11 | 84.62% |  |  |
|  | Immigrant status (> 5 years ago) | 1 | 7.69% |  |  |
|  | Immigrant status (< 5 years ago) | 1 | 7.69% |  |  |
| **Relationship Status (n=13)** | Married | 4 | 30.77% |  |  |
|  | Divorced | 1 | 7.69% |  |  |
|  | Dating a single partner | 2 | 15.38% |  |  |
|  | Not in a relationship | 6 | 46.15% |  |  |
| **Employment Status (n=13)** | Full-Time | 7 | 53.85% |  |  |
|  | Part-Time | 1 | 7.69% |  |  |
|  | Disability (not working) | 2 | 15.38% |  |  |
|  | Unemployed + volunteer work | 2 | 15.38% |  |  |
|  | Unemployed | 1 | 7.69% |  |  |
| **Highest Level of Education (n=12)** | Master's degree | 3 | 25.00% |  |  |
|  | Bachelor's degree | 1 | 8.33% |  |  |
|  | University, no degree | 3 | 25.00% |  |  |
|  | CEGEP | 3 | 25.00% |  |  |
|  | High school or equivalent (GED) | 2 | 16.67% |  |  |
| **Years of Education (n=12)** | N/A | 12 | N/A | 15 | 6.42 |
| **Income (\$ CA) (n=10)** | N/A | 10 | N/A | 82333.33 | 70099.93 |

The patient’s next appointment with their physician served as the baseline visit, where the tool was not used by the physician during the appointment. The tool was used during at least two of the subsequent appointments, which served as visits 1 and 2 (respectively). Patients were considered to have completed the study if they attended the baseline and visit 1 and 2 appointments at a minimum; completion of study related tasks was not a criteria for study completion given that this was a feasibility study focused on determining what patients would realistically complete. Research personnel recorded whether the baseline visit was an intake visit or a follow-up visit; this was relevant as initial intake visits are generally longer than follow-ups; tracking this allowed for the adjustment of analyses such that initial intake visits would not artificially inflate the visit length at baseline. In the week preceding visit 1, patients completed the Quick Inventory of Depressive Symptomatology (QIDS) and met with study personnel to be administered the Inventory of Depressive Symptomatology, Clinician Rating (IDS-C) (Yeung et al., 2012; Rush et al., 2000). These questionnaires were part of the set of questions used to generate the AI results.

Due to COVID-19 and the public health recommendations released by the Québec Government in March 2020, the study was adapted to be completed entirely from a distance. Originally, the protocol intended for the appointment length to be recorded by research personnel on-site, from the moment the patient entered the room to when they left. However, due to the transition from in-person to telemedicine (phone and video appointments), appointment length was measured as the length of the telemedicine or phone call during which the visit took place, as displayed on the physician or patient device and relayed verbally to research personnel. Further information about adaptation to COVID-19 can be found in the Supplementary Materials.

After each visit where the tool was used, physicians were asked to complete a post-appointment questionnaire describing device useability and any serious adverse events (SAE) and to use the Udvalg for Kliniske Undersøgelser Side Effects Rating Scale (UKU) (Lingjaerde et al. 1987) to record any side effects as perceived by the treating physician. Research personnel also administered the Brief Adherence Rating Scale (BARS) to patients to estimate medication adherence since the prior visit (Byerly et al. 2008).

Following visit 2, patients met with research personnel for end of study tasks, which consisted of completing the QIDS, the Scale to Assess Therapeutic Relationships in Community Mental Health Care (STAR-P) (McGuire-Snieckus, McCabe, Catty, Hansson & Priebe, 2007), a customized exit questionnaire designed specifically to capture elements of the experience of using this novel tool, as well as being administered the IDS-C and a custom semi-structured interview. After all their patients completed the study, physicians were also administered the STAR-C as well as a customized exit questionnaire designed specifically to capture elements of the experience of using this novel tool, as well as a custom semi-structured interview.

## Statistical Analyses

Appointment length at baseline, visit 1 and visit 2 was extracted from the research assistant appointment log and analyzed using Statistical Package for Social Sciences version 27 (SPSS 27) repeated measures ANOVA (three factors, within-subjects variables). Results relating to hypotheses two, three and four were derived from the exit questionnaires and were reported as means (including SD) or percentages. Additionally, we measured the percentage of questionnaires actually sent that were completed each week for the PHQ-9, GAD-7, PRISE-20 and FIBSER.

Questionnaire completion was measured from the date their account was created (week 1 in the study) up to week 12 (this timeframe was chosen because 12 weeks is the follow-up time planned for our clinical trial). For other timeframes, please see the Supplementary Material. The number of PHQ-9 questionnaires that were completed in the app by patients was calculated by subtracting those completed by physicians and taking the mean completion rate across all patients in each of the three time intervals. Note that only the PHQ-9 could be completed by physicians; all other questionnaires in the CDSS could be completed by patients only.

## Results

Ten physicians were initially recruited; however, 3 psychiatrists were unable to recruit patients due to COVID-19 related interruptions in regular clinical practice and could not be included (2 psychiatrists’ day programs were closed and 1 psychiatrist focused on providing consults rather than follow-up appointments during the pandemic). 20 patients were approached by the 10 physicians recruited for the study. Of these, 2 declined participation after discussing the study with their physician or a research assistant. One patient who was interested in entering the study was not eligible as another physician involved in prescribing their medication was not a study physician and as such would not be able to use the application to follow the patient. The recruiting physician was running a day hospital program which the patient in question was attending. This left 17 patients who were recruited into the study. As such, 85% of patients approached were recruited. 14 patients (82%) (Table 3) completed the study (defined as attending baseline, visit 1 and visit 2 appointments). 1 patient withdrew prior to the baseline appointment and 2 withdrew after the baseline appointment but before the CDSS was used at visit 1. The sample of patients completing the study consisted of 9 women and 5 men with a mean age of 36.43 (SD=±14.84). See table 3 for demographics and table 1 for baseline questionnaire scores. The pandemic also resulted in significantly reduced recruitment, and was a reason for withdrawal for several of the patients who withdrew from the study.

Two patients (11.76%) experienced side effects, as recorded on the UKU across the course of the study; please see supplementary materials for more details. One patient (5.88%) discontinued treatment. We note that discontinuation rates of psychiatric treatment can often be greater than 40% (Maund et al., 2019). There were no serious adverse events related to the tool; however, 1 patient experienced 2 emergency room visits (a work-related injury and a rash which was thought to be viral by consultants in the E.R. but which may have been related to a new antidepressant prescription that was made by a physician without reference to the AI predictions) during the study.

The average number of weeks that patients (n=14) were in the study was 13.2 weeks or 92.4 days (stdev=9.74 weeks or stdev=68.18 days), excluding two patients who dropped out of the study within one week of creating their accounts. The time between baseline appointment and visit 1 was a mean of 40.86 days (SD=29.40), and the time between visit 1 and visit 2 was a mean of 51.57 days (SD=62.58). At baseline, visits lasted a mean of 19.29 minutes (SD = 5.75). Visit 1 and Visit 2 lasted a mean of 17.80 minutes (SD = 9.74) and 21.39 minutes (SD = 10.28), respectively (Figure 2). Our findings show no significant difference between the baseline appointment time without the CDSS, and subsequent visits using the CDSS (F(2,24)=0.805, MSE=58.08, p=0.46).

With respect to the subjective physician view of appointment length, 4 of the 7 physicians rated that using the tool “took about the same time as my usual practice”, indicated by a rating of 3 using a 5-point Likert scale. Additionally, 61.54% of patients felt that their appointment time did not change, while 1 patient felt that it decreased.

With respect to the tool’s trustworthiness, 61.54% of the patients and 71.43% of the physicians rated that they trusted the CDSS, indicated by a 4 or 5 on the 5-point Likert scale. Overall usability of the CDSS, indicated by a 4 or 5, was rated 92.31% by patients and 71.43% by physicians (see Supplementary Material Table 4).

At each patients’ visit 1, 100% of physicians logged into the tool, and the clinical algorithm module (which contains the CANMAT guidelines and AI results, when available) portion of the tool was accessed at 13 out of 14 appointments. At subsequent Visit 2 appointments, once again 100% of physicians logged into the tool, while the clinical algorithm component was again accessed at 13 out of 14 appointments.

The mean STAR-P and STAR-C scores were 33.62 ± 2.90 and 31.14 ± 2.63, respectively, compared to 38.4 ± 12.0 and 31.5 ± 6.9 in the original STAR study (McGuire-Snieckus et al., 2007). Further information about the STAR subscales are present in the supplementary material. In addition, on their custom exit questionnaire 46.15% of patients felt that the patient-clinician relationship significantly or somewhat improved, while the other 53.85% felt that it did not change.

Figure 1 demonstrates the PHQ-9 and GAD-7 completion rates each week during the first 12 weeks in the study. The light bars in Panels A and B reflect the total number of questionnaires sent, given the number of patients that were active in the study during each of weeks 1 through 12. The total number of PHQ-9 and GAD-7 questionnaires completed by patients on the application for the first 12 weeks of the study were summed and are shown in the dark bars in Panels A and B. On each of weeks 4, 5, 6, and 10, one patient completed their PHQ-9 questionnaire with a physician. For each of these weeks, one response was subtracted from the total number of PHQ-9 questionnaires completed to reflect only those done by patients alone. See Figure 1, Panel C and D.

**Figure 1.**
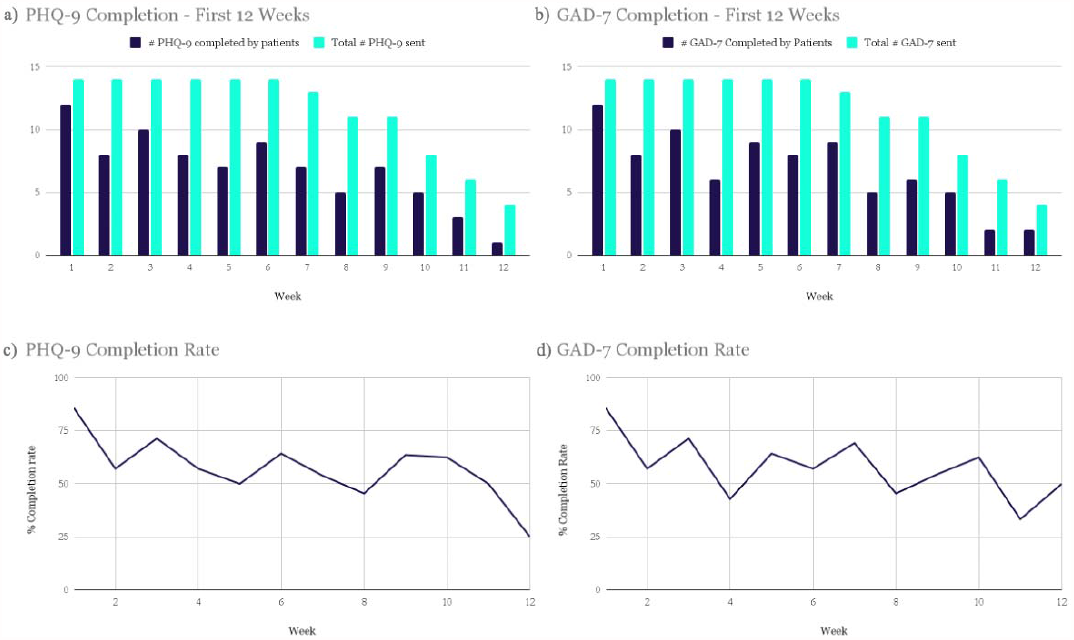
Panel A demonstrates the number of PHQ-9 questionnaires completed by patients in the first 12 weeks of the study versus the total number sent in the CDSS (1 per week, per active patient); Panel B demonstrates the number of GAD-7 questionnaires completed by patients in the first 12 weeks of the study versus the total number sent in the CDSS (1 per week, per active patient); Panel C demonstrates the percent of PHQ-9 questionnaires that were completed by patients in the CDSS during the first 12 weeks of the study; and Panel D demonstrates the percent of GAD-7 questionnaires that were completed by patients in the CDSS during the first 12 weeks of the study.

**Figure 2.**
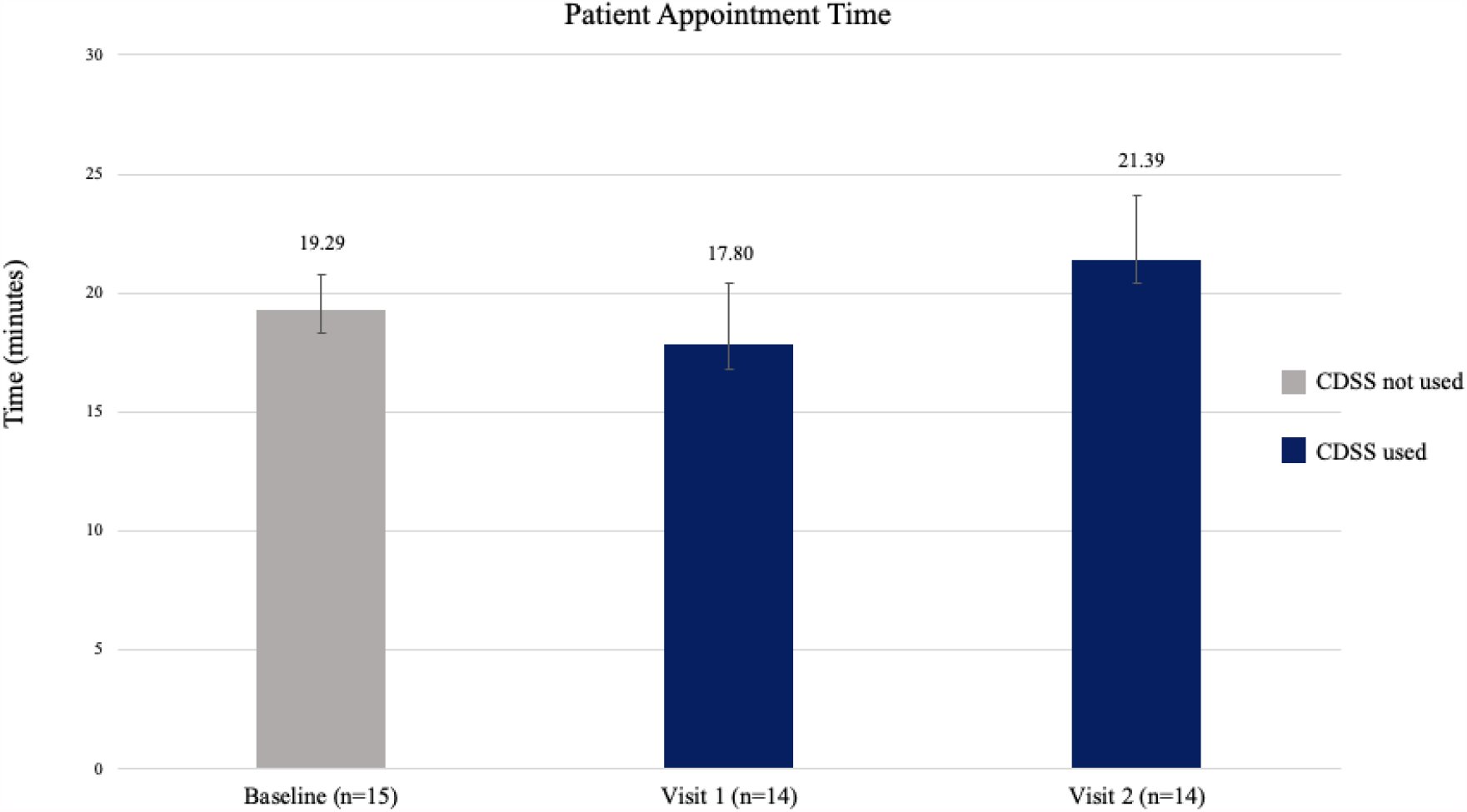
Patient appointment times. 16 patients were assessed at the baseline appointment; however, 1 patient’s appointment time could not be counted at baseline due to their physician opening the CDSS in error.

The mean completion rate of all PHQ-9 questionnaires sent from account creation to week 12 of the study was 64%. The mean completion rate of PHQ-9 by patients alone in this time frame was 59%. The GAD-7 had a mean completion rate of 60% in the first 12 weeks of the study. Completion rates for other time frames can be found in the Supplementary Materials but were similar.

Of the participants (n=14), 10 had mean PHQ-9 and GAD-7 completion rates equal to or greater than 50%. The lowest completion rate among the patients for both questionnaires sent in the first 12 weeks (n=1) was 8%. Most patients regularly completed questionnaires, with 76% (n=11) completing ≥ 33% of app assessments and 71% (n=10) completing ≥ 50% of PHQ-9 and GAD-7 assessments in the app over the first 12 weeks in the study.

Bi-weekly peaks in the completion rates were observed for the PHQ-9 and GAD-7 questionnaires (Figure 1, Panel C and D). Although they were intended to be completed every week as a part of the study, these questionnaires are often administered at two week or greater intervals in practice, indicating the likely feasibility of a reduced questionnaire frequency.

In addition, we found in an exploratory analysis aimed at determining potential correlates of questionnaire completion rates that patients who had appointments scheduled further apart were less likely to complete the PHQ-9 (*r*(12)= -0.69, *p* = 0.0063).

Results and a full discussion of the changes in depression and anxiety questionnaire scores can be found in the supplementary material.

## Discussion

The primary objectives of the study were to assess feasibility, useability and ongoing engagement with the CDSS when integrated into clinical practice, as well as to measure physician and patient trust in the CDSS. The primary outcome of interest was session length, in order to determine whether the use of the CDSS required more time than the baseline appointment. We were able to confirm our primary hypothesis that session length did not increase after the introduction of the tool. Moreover, it was crucial to ensure that the tool did not impose an undue burden on either the physician or the patient throughout the duration of the study.

Digital application use for the purpose of promoting mental health is increasingly recommended by public health organizations. For example, in its Mental Health Action Plan 2013–2020, the World Health Organization proposed “the promotion of self-care, for instance, through the use of electronic and mobile health technologies’’ (Anthes, 2016). Meanwhile, the UK National Health Service (NHS)’s website endorses a short list of online mental-health resources, which includes smartphone-based apps (Anthes, 2016). Specifically, studies on app usability in the treatment of depression demonstrate that telemedicine and internet-based approaches are feasible and as effective as in-person treatment (Arean et al., 2016).

However, these tools also face substantial barriers with regard to adherence. For example, a randomized clinical field trial conducted by Arean et al. compared three different mobile apps for depression, in order to examine how individuals who download these tools typically use them (Arean et al., 2016). The authors’ findings show that most participants did not use their assigned intervention apps as instructed, and experienced a significant drop off in usage after two weeks (Arean et al., 2016). Additionally, a study that investigated the feasibility of using a smartphone app to assess schizophrenia prompted patients via text message to complete personalized questionnaires once per week. They found that participants (n=18) completed 65% of app assessments, “with 78% completing ≥33% app assessments and 72% completing ≥50% app assessments” (Eisner et al., 2019), similar to the results observed in this study. In summary, response rates observed in our study were reasonable in the context of previous reports and engagement persisted fairly stably beyond two weeks, demonstrating the application was able to retain patients at least as or more consistently than were applications in previous studies.

Physician engagement with mental health apps is related to their technological competency, their perception of patient access to technology, and organizational infrastructure that facilitates the adoption of the apps into their practice (Chiauzzi and Newell, 2019); these were all factors considered when designing the study-for example, physicians who were less technically oriented could rely on study staff to provide ongoing support for application use as needed. The results of our study demonstrate sustained patient and physician engagement beyond two weeks, potentially due to the fact that the application was directly tied to clinical care, and because high physician usage of the application and the data patients inputted may have motivated patients to continue engaging. Indeed, higher rates of engagement are linked to the use of telephone and email reminders, as well as follow-up with a physician (Christensen et. al, 2009), a finding we support with our demonstration of lower PHQ-9 response rates as a result of longer inter-appointment lengths. In addition, the email reminders sent to patients to complete assessments likely had a positive impact on completion rates, based on these previous findings.

More than half (4 out of 7) of physicians felt that using the tool in session took approximately the same time as their usual practice. It is important to note that the second hypothesis assessed the physicians’ subjective view on appointment length, rather than the objective appointment time. Therefore, it is possible that some physicians reported that their appointments felt longer than their usual practice simply because they were not yet familiar with the tool. Interestingly, the majority of patients did not subjectively report that the tool increased their appointment time. Objectively, appointment length did not increase when the tool was introduced, lending credence to the idea that the novelty of the tool use may have influenced perception of time spent by physicians.

The third hypothesis predicted that at least 66% of patients and 66% of physicians would rate the trustworthiness of the CDSS as a 4 or 5. Physicians confirmed this hypothesis, where 71.43% rated the tool as trustworthy; patients came close, cumulatively rating trust at 61.54%; the slightly lower than expected score may be due to small sample size and the impact of COVID-19: most clinicians followed up with their patients by phone, which meant that patients did not get to view the AI results on the physician’s screen as intended, which may have improved feelings of trust had it occurred more frequently; we note that standardized patients noted that looking at the screen with their physicians was a positive experience in our previous simulation center study (Benrimoh et. al., 2020). Nonetheless, patients’ mean score was 3.85 using the 5-point Likert scale, which indicates that patient trust trends in a positive direction.

The fourth hypothesis predicted that both physicians and patients would rate the overall useability of the tool at least 66%. The hypothesis was confirmed as 71.43% of physicians and 92.31% of patients considered the tool easy to use, again demonstrating that the tool did not impose an unwarranted burden on either patients or clinicians.

For the fifth hypothesis, we predicted that at least 65% of patients would still be using the application regularly by the end of the study. We defined “regularly” as the completion of at least one PHQ-9 and GAD-7 questionnaire on the application per week. We note that our results did not meet the threshold set by the hypothesis; however, it is likely that the hypothesis, which required a patient to have completed all of their questionnaires every week to be considered “engaged”, was overly strict and that the completion rates described above are a better metric as they are more in line with the results from previous studies. In addition, the small sample size means that a few patients with significantly worse performance than the mean could significantly impact the results. We also note that some patients were “poor completers”-that is, they did not regularly fill out questionnaires for the majority of the study; the majority of patients, however, filled out 50% or more of their assessments, as noted above. Considering this data in the context of the previous reports of patient engagement with digital tools described above, we consider the tool to have had acceptable levels of patient engagement.

Finally, we hypothesized that at least 70% of physicians would still be using the application regularly by the end of the study, with ‘regularly’ defined as application use in every study-related visit. The tool was accessed by physicians at every visit 1 and 2, demonstrating regular application use over the course of the study. The clinical algorithm component of the tool was also accessed at approximately 92% of all appointments. As such, our results surpassed the expectations set forth by this hypothesis.

## Limitations

The main limitation of the study is the small sample size and heterogeneity in terms of patient depression severity illness stage. Nevertheless, it also presents as a strength because it allowed us to demonstrate feasibility in a range of clinical situations.

A significant limitation of this study is that while its design allowed us to examine the impact of introducing the tool on the patient-clinician relationship and the clinician workflow, this at the same time prevented us from examining effectiveness of the device in terms of improvement in depression scores. This is because the tool, being introduced well into a patient’s treatment course, could not have its intended effect of assisting treatment selection or to help clinicians implement measurement-based care and algorithm-guided treatment across the entire length of the study. This was compounded by the delays between appointments and the reduced number of visits as a result of COVID-19. However, we note that the decision was made during study design not to focus on effectiveness and, due to the novelty of the device and the need to determine challenges to its introduction into clinical practice, to focus squarely on feasibility. As such, the modest improvements in depression and anxiety scores seen here are in line with expectations given that the tool was not introduced in a manner where it could have its intended effect on patient care.

In this paper, we have demonstrated that the Aifred CDSS is feasible and easy for clinicians and patients to use in a longitudinal manner, that it does not require increased time to use in clinic, and that it may have beneficial effects on the physician-patient relationship (this latter point will be further elaborated in a future article). The next step in the evaluation of this software will be a clinical trial aimed at investigating the safety and effectiveness of the CDSS.

## Data Availability

Inquiries regarding access to study data should be directed to the corresponding author.

## Data Availability

Inquiries regarding access to study data should be directed to the corresponding author.

## Data Availability

Inquiries regarding access to study data should be directed to the corresponding author.

## Acknowledgements

We thank the patients, participating doctors and local staff for being involved in this feasibility study.

## References

1. Zikos, D., DeLellis, N. CDSS-RM: A clinical decision support system reference model. BMC Med Res Methodol 18, 137 (2018). https://doi.org/10.1186/s12874-018-0587-6

2. Sutton, R.T., Pincock, D., Baumgart, D.C. et al. An overview of clinical decision support systems: benefits, risks, and strategies for success. npj Digit. Med. 3, 17 (2020). https://doi.org/10.1038/s41746-020-0221-y

3. Torous, J., Lipschitz, J., Ng, M., Firth, J. (2020, Feb) Dropout rates in clinical trials of smartphone apps for depressive symptoms: A systematic review and meta-analysis. Journal of Affective Disorders, 263, 413–419. https://doi.org/10.1016/j.jad.2019.11.167.

4. Rollman, B. L., Hanusa, B. H., Lowe, H. J., Gilbert, T., Kapoor, W. N., & Schulberg, H. C. (2002). A randomized trial using computerized decision support to improve treatment of major depression in primary care. Journal of General Internal Medicine, 17(7), 493–503.

5. Harrison, P., Carr, E., Goldsmith, K., Young, A., Ashworth, M., Fennema, D., Barrett, B., & Zahn, R. (Accepted/In press). Study Protocol for The Antidepressant Advisor (ADeSS): A Decision Support System for Antidepressant Treatment for Depression in UK primary care - a feasibility study: The Antidepressant Advisor. BMJ Open, 10:e035905.

6. Adli, M., Wiethoff, K., Baghai, T. C., Fisher, R., Seemüller, F., Laakmann, G., … & Bauer, M. (2017). How effective is algorithm-guided treatment for depressed inpatientsã Results from the randomized controlled multicenter German algorithm project 3 trial. International Journal of Neuropsychopharmacology, 20(9), 721–730.

7. Warden, D., Rush, A. J., Trivedi, M. H., Fava, M., & Wisniewski, S. R. (2007). The STAR* D Project results: a comprehensive review of findings. Current psychiatry reports, 9(6), 449–459.

8. Benrimoh, D., Tanguay-Sela, M., Perlman, K., Israel, S., Mehltretter, J., Armstrong, C., … Margolese, H. (2021). Using a simulation centre to evaluate preliminary acceptability and impact of an artificial intelligence-powered clinical decision support system for depression treatment on the physician–patient interaction. BJPsych Open, 7(1), E22. doi:10.1192/bjo.2020.127

9. Benrimoh, D., Fratila, R., Israel, S., Perlman, K., Mirchi, N., Desai, S., … & You, R. P. (2018). Aifred health, a deep learning powered clinical decision support system for mental health. In The NIPS’17 Competition: Building Intelligent Systems (pp. 251–287).

10. Kennedy, S. H., Lam, R. W., McIntyre, R. S., Tourjman, S. V., Bhat, V., Blier, P., Hasnain, M., Jollant, F., Levitt, A. J., MacQueen, G. M., McInerney, S. J., McIntosh, D., Milev, R. V., Müller, D. J., Parikh, S. V., Pearson, N. L., Ravindran, A. V., Uher, R., & CANMAT Depression Work Group (2016). Canadian Network for Mood and Anxiety Treatments (CANMAT) 2016 Clinical Guidelines for the Management of Adults with Major Depressive Disorder: Section 3. Pharmacological Treatments. Canadian journal of psychiatry. Revue canadienne de psychiatrie, 61(9), 540–560. https://doi.org/10.1177/0706743716659417

11. Ventola C. L. (2014). Mobile devices and apps for health care professionals: uses and benefits. P & T : a peer-reviewed journal for formulary management, 39(5), 356–364.

12. Canadian Institutes of Health Research, Natural Sciences and Engineering Research Council of Canada, and Social Sciences and Humanities Research Council (2005), Tri-Council Policy Statement: Ethical Conduct for Research Involving Humans.

13. American Psychiatric Association. (2013). Diagnostic and statistical manual of mental disorders (5th ed.). https://doi.org/10.1176/appi.books.9780890425596

14. Kroenke, K., Spitzer, R. L., & Williams, J. B. (2001). The PHQ-9: validity of a brief depression severity measure. J Gen Intern Med, 16(9), 606–613. https://doi.org/10.1046/

15. Spitzer, R. L., Kroenke, K., Williams, J. B., & Löwe, B. (2006). A brief measure for assessing generalized anxiety disorder: the GAD-7. Arch Intern Med, 166(10), 1092–1097. https://doi.org/10.1001/archinte.166.10.1092

16. Bohn, M. J., Babor, T. F., & Kranzler, H. R. (1995, Jul). The Alcohol Use Disorders Identification Test (AUDIT): validation of a screening instrument for use in medical settings. J Stud Alcohol, 56(4), 423–432. https://doi.org/10.15288/jsa.1995.56.423

17. Merlhiot G, Mondillon L, Vermeulen N, Basu A, Mermillod M (2014) Adaptation and Validation of the Standardized Assessment of Personality – Abbreviated Scale as a Self-Administered Screening Test (SA-SAPAS). J Psychol Psychother 4: 164. https://doi.org/10.4172/2161-0487.1000164

18. Wisniewski, S. R., Rush, A. J., Balasubramani, G. K., Trivedi, M. H., & Nierenberg, A. A. (2006). Self-rated global measure of the frequency, intensity, and burden of side effects. J Psychiatr Pract, 12(2), 71–79. https://doi.org/10.1097/00131746-200603000-00002

19. Felitti, V. J., Anda, R. F., Nordenberg, D., Williamson, D. F., Spitz, A. M., Edwards, V., Koss, M. P., & Marks, J. S. (1998). Relationship of childhood abuse and household dysfunction to many of the leading causes of death in adults. The Adverse Childhood Experiences (ACE) Study. American journal of preventive medicine, 14(4), 245–258. https://doi.org/10.1016/s0749-3797(98)00017-8

20. Weathers, F.W., Blake, D.D., Schnurr, P.P., Kaloupek, D.G., Marx, B.P., & Keane, T.M. (2013). The Life Events Checklist for DSM-5 (LEC-5).

21. Yeung, A., Feldman, G., Pedrelli, P., Hails, K., Fava, M., Reyes, T., & Mundt, J. C. (2012). The Quick Inventory of Depressive Symptomatology, clinician rated and self-report: a psychometric assessment in Chinese Americans with major depressive disorder. The Journal of nervous and mental disease, 200(8), 712–715.

22. Rush, A.J., Carmody, T. and Reimitz, PLJE. (2000), The Inventory of Depressive Symptomatology (IDS): Clinician (IDSLJC) and SelfLJReport (IDSLJSR) ratings of depressive symptoms. Int. J. Methods Psychiatr. Res., 9: 45–59.

23. Lingjaerde, O., Ahlfors, U. G., Bech, P., Dencker, S. J., & Elgen, K. (1987). The UKU side effect rating scale. A new comprehensive rating scale for psychotropic drugs and a cross-sectional study of side effects in neuroleptic-treated patients. Acta Psychiatr Scand Suppl, 334, 1–100. https://doi.org/10.1111/j.1600-0447.1987.tb10566.x

24. Byerly, M. J., Nakonezny, P. A., & Rush, A. J. (2008, Mar). The Brief Adherence Rating Scale (BARS) validated against electronic monitoring in assessing the antipsychotic medication adherence of outpatients with schizophrenia and schizoaffective disorder. Schizophr Res, 100(1-3), 60–69. https://doi.org/10.1016/j.schres.2007.12.470

25. Edwards, I. R., & Aronson, J. K. (2000). Adverse drug reactions: definitions, diagnosis, and management. The Lancet, 356(9237), 1255–1259.

26. McGuire-Snieckus, R., McCabe, R., Catty, J., Hansson, L., & Priebe, S. (2007, Jan). A new scale to assess the therapeutic relationship in community mental health care: STAR. Psychol Med, 37(1), 85–95. https://doi.org/10.1017/s0033291706009299

27. Gaynes, B. N., Warden, D., Trivedi, M. H., Wisniewski, S. R., Fava, M., & Rush, A. J. (2009). What did STAR*D teach usã Results from a largeLJscale, practical, clinical trial for patients with depression. Psychiatric Services, 60(11), 1439– 1445.

28. Williams, N. (2014). The GAD-7 questionnaire (Questionnaire review). Occupational Medicine, 64(3), 224–224

29. Kroenke K. (2012). Enhancing the clinical utility of depression screening. CMAJ : Canadian Medical Association journal = journal de l’Association medicale canadienne, 184(3), 281–282.

30. Toussaint, A., Hüsing, P., Gumz, A., Wingenfeld, K., Härter, M., Schramm, E., & Löwe, B. (2020). Sensitivity to change and minimal clinically important difference of the 7-item Generalized Anxiety Disorder Questionnaire (GAD-7). Journal of affective disorders, 265, 395–401.

31. Canadian Agency for Drugs and Technologies in Health (2016). Common Drug Review, Clinical Review Report, Aripiprazole (Abilify), 45.

32. Fortney, J. C., Unützer, J., Wrenn, G., Pyne, J. M., Smith, G. R., Schoenbaum, M., & Harbin, H. T. (2017). A tipping point for measurement-based care. Psychiatric Services, 68(2), 179–188.

33. Anthes, E. (2016). Mental health: there’s an app for that. Nature News, 532(7597), 20.

34. Arean, P. A., Hallgren, K. A., Jordan, J. T., Gazzaley, A., Atkins, D. C., Heagerty, P. J., & Anguera, J. A. (2016). The use and effectiveness of mobile apps for depression: results from a fully remote clinical trial. Journal of medical Internet research, 18(12), e330.

35. Zhang, R., Nicholas, J., Knapp, A. A., Graham, A. K., Gray, E., Kwasny, M. J., … & Mohr, D. C. (2019). Clinically meaningful use of mental health apps and its effects on depression: mixed methods study. Journal of medical Internet research, 21(12), e15644.

36. Li I, Dey A, Forlizzi J. A stage-based model of personal informatics systems. 2010 Presented at: SIGCHI Conference on Human Factors in Computing Systems (CHI 10); April 10-15, 2010; Atlanta, GA, USA p. 10–15. doi: 10.1145/1753326.1753409

37. Eisner, E., Bucci, S., Berry, N., Emsley, R., Barrowclough, C., & Drake, R. J. (2019). Feasibility of using a smartphone app to assess early signs, basic symptoms and psychotic symptoms over six months: A preliminary report. Schizophrenia research, 208, 105–113.

38. Christensen, H., Griffiths, K. M., & Farrer, L. (2009). Adherence in internet interventions for anxiety and depression. Journal of medical Internet research, 11(2), e13.

39. Chiauzzi, E., & Newell, A. (2019). Mental Health Apps in Psychiatric Treatment: A Patient Perspective on Real World Technology Usage. JMIR mental health, 6(4), e12292.

40. Maund, E., Stuart, B., Moore, M., Dowrick, C., Geraghty, A. W., Dawson, S., & Kendrick, T. (2019). Managing antidepressant discontinuation: a systematic review. The Annals of Family Medicine, 17(1), 52–60.

41. Cipriani, A., Furukawa, T. A., Salanti, G., Chaimani, A., Atkinson, L. Z., Ogawa, Y., … & Geddes, J. R. (2018). Comparative efficacy and acceptability of 21 antidepressant drugs for the acute treatment of adults with major depressive disorder: a systematic review and network meta-analysis. Focus, 16(4), 420–429.

42. Mehltretter, J., Fratila, R., Benrimoh, D., Kapelner, A., Perlman, K., Snook, E., … & Turecki, G. (2020). Differential treatment benefit prediction for treatment selection in depression: a deep learning analysis of STAR* D and CO-MED data. Computational Psychiatry, 4, 61–75.

43. Mehltretter, J., Rollins, C., Benrimoh, D., Fratila, R., Perlman, K., Israel, S., … & Turecki, G. (2020). Analysis of features selected by a deep learning model for differential treatment selection in depression. Frontiers in Artificial Intelligence, 2, 31.

44. Squarcina, L., Villa, F. M., Nobile, M., Grisanc, E., & Brambilla, P. (2020). Deep learning for the prediction of treatment response in depression. Journal of Affective Disorders.

45. Pew Research Center. (2020). U.S. Public Sees Multiple Threats From the Coronavirus – and Concerns Are Growing. https://www.pewresearch.org/politics/2020/03/18/u-s-public-sees-multiple-threats-from-the-coronavirus-and-concerns-are-growing/

