## Supplementary Material for "Evaluating the Clinical Feasibility of an Artificial Intelligence-Powered Clinical Decision Support System: A Longitudinal Feasibility Study"

SIMULATION STUDY BACKGROUND:

In a previous simulation centre study, we evaluated the utility of the CDSS as perceived by primary care physicians and psychiatrists participating in simulated clinical interactions with standardized patients (Benrimoh et al., 2020). 60% of physicians perceived the CDSS to be a useful tool in their treatment selection process for major depressive disorder (MDD) (Benrimoh et al., 2021). Furthermore, 50% of physicians reported they would use the tool for all depression patients, with an additional 35% reporting they would reserve tool use for more severe or treatment-resistant patients (Benrimoh et al., 2021). In addition, during the study, we were able to determine that the CDSS, as designed, could be integrated into a clinical interview in a short time (< 5 minutes if needed) and that it did not have significant negative effects on the patient-clinician interaction.

TELEMEDICINE DURING COVID-19

Research personnel administered questionnaires and interviews over the phone with patients and physicians. Self-report questionnaires and consent forms were sent to patients and physicians via email, who were instructed to keep completed versions of these documents in a secure location at home or their office, in either a digitized or print form; these were then later collected by research staff when it was safe to do so. Both patients and physicians were administered a COVID-19 Anxiety questionnaire, adapted from Pew Research 2020, to assess the perceived impact of the pandemic on their individual as well as societal wellbeing (Pew Research Center, 2020). Refer to Tables 1 and 2 in supplementary for results.

PATIENT CHARACTERISTICS:

 10 of whom were of Caucasian ethnicity and had post-secondary education. 7 were working full-time. Patients (n=15) had a mean initial PHQ-9 score of 14.80 (SD=5.61). Of 15 patients, 10 had a positive screen for a personality disorder using the SAPAS-SR, 3 were engaging in harmful amounts of alcohol consumption as per the AUDIT and 4 were engaging in a substantial or severe level of drug abuse as per the DAST-10.

PATIENT MEDICATIONS:

All medications that were recorded in the CDSS were cross-referenced with the research assistant log and demographics questionnaire to understand which medications were active, newly prescribed or stopped at visit 1, visit 2, and visit 3 or 4, for those who had appointments. Medications were omitted in the data analysis if 1) there was an error (e.g., the clinician or patient entered accidental prescriptions) or if 2) physicians prescribed and ended the prescription on the same appointment date. Our research looked to investigate the total number of medications prescribed and discontinued throughout the study and the average number of depression medications for all patients.

Patients (n=14) had a mean of 2.29 medications (SD=1.20) throughout the study, where 9 new medications were prescribed, 2 medications were stopped and 8 dosages were changed. There were 3 medication changes between visit 1 and visit 2; 1 was a dosage change and 2 were medications added. Upon study close-out, 28 medications were active across patients. As such, there were very few medication changes which the tool could have had an effect.

Many patients (10/14) entered the study prescribed medications that persisted past visit 1. One patient was not prescribed any medication but instead was referred to therapy given that their depression was not of a severity where their clinician found that medication was warranted. Throughout the study, the most common medication was Bupropion XL, followed by Citalopram and Trazodone. 64.29% (9/14) of medication plans were changed, either due to changing the dosage, adding or discontinuing a medication. This is understandable given that clinicians often wait at least 8 weeks after starting a treatment before making a change (Warden et al., 2007). Most physicians altered treatment plans during appointments with patients; however, one physician changed the treatment plan three times between visit 1 and visit 2.

MACHINE LEARNING PREDICTIONS:

Machine learning (ML) remission probabilities were generated at visit 1 and visit 2 for patients who had completed QIDS, IDS-C and CQ. We looked to investigate how many medications were prescribed during visit 1 and visit 2, of the ML predictions that were seen by physicians. Additionally, we were interested in identifying the ML remission probability ranking compared to the patient’s treatment plan, regardless of whether the physician saw the ML prediction. Lastly, we examined the completion date for each questionnaire used to generate ML predictions at visit 1 and visit 2, so that we could see to see how many days passed between the majority of questionnaires being completed and the appointment.

Before visit 1, physicians saw the ML predictions for 7 patients. Of the 7 generated predictions, 2 patients were prescribed a medication from the ML prompted remission probability list. Both medications were the fourth-highest remission probability. Before visit 2, physicians saw the ML predictions for 10 patients. Of the 10 predictions, 1 patient was prescribed a medication from the ML results, with it producing the 7th highest remission probability. An 11th patient’s ML prediction was generated after their visit 2 appointment and thus, the physician never saw the results. At visit 1, 5/7 patients completed the three questionnaires on the same day, where 2 patients completed their third questionnaire 2 days and 29 days later, respectively. The average time between questionnaire completion and visit 1 was 9.43 days (SD=11.25). At visit 2, 5/10 patients completed the three questionnaires on the same day, where 5 patients completed the third questionnaire 1 or 2 days before or after the other questionnaires.  The average time between questionnaire completion and visit 2 was 66.50 days (SD=81.76).

Despite only a few patients being prescribed the medication that was recommended by the ML prediction, it is worth noting that 4 medications were already being taken by patients prior to visit 1 that matched suggestions produced by the ML.  Similarly, 10 medications that corresponded with ML predictions at visit 2 were already in the patient's treatment plans. Most patients completed the three ML-required questionnaires on the same day. Understandably, the days between questionnaire completion and appointments increased considerably between visit 1 and visit 2, most likely due to the fact that patients completed the required questionnaires near the beginning of the study and therefore, these were used to generate ML predictions at visit 2, despite their answers being inputted months earlier.

REMISSION AND RESPONSE CUT-OFFS:

 Remission (the clinical response of greatest interest) is defined as a PHQ-9 score < 5 at 12 months, a QIDS score ≤ 5 (Yeung et al., 2012), and an IDS-C score ≤ 13 (Rush et al., 2000).  Response was defined as patients who saw a ≥ 50% decrease in questionnaire scores. A clinically significant improvement on the PHQ-9 is defined as having a score less than 10 with a 50% decrease from the pre-treatment score (Kroenke et al., 2001). There is no remission rate for the GAD-7, however a score ≤ 5 represents low to no anxiety (Williams, 2014). The minimum clinically important difference (MCID) is defined as the minimum change in questionnaire score from baseline in which the patient perceives a difference as a result of treatment or intervention. This represents a 5-point decrease on the PHQ-9 (Kroenke, 2012) and a 4-point decrease on the GAD-7 (Toussiant et. al, 2020). No MCID has been specified for either the QIDS or IDS-C (CADTH, 2016).

Remission (the primary exploratory depression endpoint of interest) was observed for 28.6% (n=4) of patients based on PHQ-9 scores; 14.3% (n=2) of patients reached remission based on QIDS scores and 14.3% (n=2) of patients reached remission based on IDS-C scores. Response was observed for 28.6% (n=4) of patients based on PHQ-9 scores; 7.1% (n=1) of patients based on QIDS scores; 21.4% (n=3) of patients as reported by IDS-C scores; and 42.9% (n=6) of patients based on GAD-7 scores. The lower rates of response and remission on the QIDS and IDS-C may have been affected by a lack of complete data for these questionnaires. There were 5 patients who did not complete either the first, last or both QIDS questionnaires and 2 patients did not complete either the first, last or both IDS-C questionnaires. The decrease in scores across the PHQ-9, GAD-7, QIDS and IDS-C questionnaires suggests that overall, patients saw modest improvements; however, none of the findings were statistically significant (see the supplementary section Questionnaire Score Changes).

APPOINTMENT LENGTH AND SCHEDULING:

Results confirmed our primary hypothesis that the CDSS would not significantly increase visit length when incorporated into clinical practice. Due to the implications of the COVID-19 pandemic, in-person appointments transitioned to telemedicine and follow-ups via phone. Both physicians and patients had access to the CDSS during these telemedicine appointments. Furthermore, appointments were not scheduled as frequently as initially intended. Initially, these were supposed to occur at least once per month; however, this proved impossible amidst the reorganization caused by the pandemic. As a result, clinicians scheduled patients when they were able to do so; longer delays, as we noted, were correlated with lower questionnaire response rates. Potentially, patients were less inclined to complete questionnaires because they knew their physician would not see them until weeks later. Previous work has noted that in order for measurement-based care to be effective and engage patients, patients must be assessed frequently and shortly before or after appointments (Fortney et al., 2017).

QUESTIONNAIRE COMPLETION:

Questionnaire completion was measured across three time frames: from the date their account was created (week 1 in the study) up to week 12 (this timeframe was chosen because 12 weeks is the follow-up time planned for our clinical trial); between account creation and visit 2 (which represents the full time a patient had access to the application); and between baseline and visit 2 (representing the time the patient was seeing their clinician during the study). We chose three time frames to better understand patient response patterns in different phases of the project.

PHQ-9 COMPLETION RATE:

The total completion rate for the PHQ-9 was 62% from account creation to visit 2. The total completion rate for the PHQ-9 was 63% from baseline to visit 2. The PHQ-9 completion rate for those completed only by patients from account creation to visit 2, was 58%. The completion rate was also 58% from baseline to visit 2.

GAD-7 COMPLETION RATE:

The GAD-7 had a mean completion rate of 60% in the first 12 weeks of the study. From account creation to visit 2 the completion rate was 58%. The completion rate was also 58% from baseline to visit 2.

STAR SUBSCALES:

The STAR comprises a clinician version (STAR-C) and a patient version (STAR-P), each with 12 items evaluated on a 5-point scale ranging from 0 to 4. Total scores may range from 0 to 48, where a higher score indicates a more favourable therapeutic relationship. We did not administer the STAR at the beginning of the study to explore the change in therapeutic relationships throughout the study period, since patients began their participation having different relationship durations and qualities with their physician that we could not control for due to the small sample size.

The mean STAR-P subscores for positive collaboration, positive clinical input and non-supportive clinician input were 21.92 ± 2.24, 10.23 ± 1.88 and 10.54 ± 2.58, respectively, compared to 19.9 ± 6.7, 9.3 ± 3.0 and 9.3 ± 3.3 in the original STAR study. The mean STAR-C subscores for positive collaboration, emotional difficulties and positive clinician input were 17 ± 2.60, 10.57 ± 1.79 and 9.93 ± 1.38, respectively, compared to 15.3 ± 4.0, 7.4 ± 2.7 and 8.9 ± 1.6 in the original STAR study. Total STAR-P scores had a correlation of 0.42790 (p=0.1447) with total STAR-C scores, 0.21705 (p=0.4763) with patient’s mean inter-appointment length, and -0.25885 (p=0.3931) with patient’s perceived effect of using the tool on their relationship with their physician.

FIBSER:

The Frequency, Intensity, and Burden of Side Effects Rating (FIBSER) questionnaire was used to measure side effects in the patient sample group. The questionnaire uses three questions with a 6-point Likert scale to measure three side-effect domains of impact: frequency, intensity and burden. A score of 0-2 on question 3 (burden of side effects) represents an acceptable low side-effect burden usually requiring no treatment adjustment. A score of 3 or 4 indicates a moderate side-effect burden that should be evaluated further and an adjustment such as a dosage decrease considered. A score of 5 to 6 indicates a high burden warranting a change in dosage or direct treatment of the side effects. (Wisniewski et al.,)

79% (n=11) of the sample group reported that they experience side effects less than 25% of the time as a result of antidepressant medications. 14% (n=2) reported they experience side effects 26-75% of the time and 7% (n=1) reported they experience side effects 76-100% of the time as a result of medications. 4 participants reported a decrease in the amount of time they experience side effects throughout the duration of the study. 2 participants reported an increase in the amount of time they experience side effects throughout the duration of the study and 7 reported no change.

71% (n=10) of the sample group reported that the severity of their side effects was mild or non-existent (score of 0-2). 29% (n=4) of the sample group reported side effects of moderate to marked severity (score of 3 or 4). 6 participants reported a decrease in side effect severity throughout the duration of the study. 3 participants reported an increase in side effect severity during the study and 5 reported no change.

86% (n=12) of the sample group reported that side effects from antidepressant medications had none to mild interference with day-to-day activities (score of 0-2). 14% (n=2) of the sample group reported that side effects from antidepressant medication had moderate to marked interference with day-to-day activities (score of 3 or 4). No participants reported that side effects resulting from antidepressant medications had a severe interference with day-to-day activities (score of 5 or 6). 4 participants reported an increase in how much side effects interfered with day-to-day activities throughout the duration of the study. 4 participants reported a decrease in how much side effects interfered with day-to-day activities, and 6 participants reported no change.

The FIBSER had a mean completion rate of 51% in both the first 12 weeks of the study and from account creation to visit 2, and 50% from account creation to visit 2.

PRISE:

The Patient Related Inventory of Side Effects (PRISE) questionnaire is a patient self-report used to qualify side effects of antidepressants by identifying and evaluating the tolerability of twenty different symptoms. The patient rates whether or not symptoms are tolerable or distressing over a seven-day period. A higher score indicates more distressing side effects from antidepressants, and a lower score indicates more tolerable or no side effects. 5 patients had a higher total score during their last week of the study than at baseline. 9 patients had a lower total score during their last week of the study compared to their baseline score, and 1 patient reported no change. The completion rate for the PRISE-20 was 58% for both the first 12 weeks in the study and from baseline to visit 2 but had a lower completion rate of 52% from account creation to visit 2.

ADVERSE EVENTS:

 There were 5 AEs experienced by 2 patients, as reported by their physicians. One patient experienced lassitude, tension/inner unrest, reduced duration of sleep, tremor, palpitations/ tachycardia, weight gain, and diminished sexual desire. Another patient experienced concentration difficulties, lassitude, sleepiness, depression, tension/inner unrest, increased and reduced duration of sleep, emotional indifference, akathisia, nausea/vomiting, constipation, increased tendency to sweating, urticaria, weight gain, diminished sexual desire, tension headache and migraine; causing the discontinuation of the current treatment.

TOOL USE:

For the three patients who had a Visit 3 appointment with their physician, the tool was used 67% of the time. At each of the two appointments where the tool was used, the clinical algorithm was also accessed. One patient had a final Visit 4 with their physician, at which time the tool was not used.

QUESTIONNAIRE SCORE CHANGES:

Questionnaire score changes were calculated by taking the difference between scores obtained prior to the baseline visit and after visit 2. In addition to raw score changes, remission, response, and minimum clinically important difference rates were calculated in an exploratory manner. We note that as patients had begun treatment prior to clinician use of the tool, which was not used at baseline, by design the study is not meant to investigate the effect of the CDSS on treatment effectiveness and all clinical outcome measurements are exploratory and were not intended to reflect the effectiveness of the tool in improving treatment outcomes. This was done in order to have a design in which we could examine the effects of the introduction of the tool into clinical practice.  In addition, delays caused by the COVID-19 pandemic meant that many patients spent longer than expected without seeing their clinician between appointments, which likely affected the changes on questionnaire scores. We note that this study was not powered to detect changes in questionnaire scores and that patients were at various phases of their treatment at study start, with some patients being chronic in nature and others experiencing recent-onset depression, and with patients generally having begun their treatment course. There was also variability in the severity of the depression at baseline. This heterogeneity was valuable from a feasibility standpoint as it allowed us to test the tool in various contexts and types of patients, but it does make it more challenging to investigate treatment effects.

The average questionnaire score for the PHQ-9 was 14.80 (SD=5.61) at baseline and 11.21 (SD=6.73) at visit 2, reflecting a mean score change of -3.59 (SD=7.15) during the study. This indicates the absence of MCID. The average questionnaire score for the GAD-7 was 12.14 (SD=5.78) at baseline and 8.07 (SD=6.55) at visit 2, reflecting a mean score change of -4.21 (SD=5.89) during the study, which meets criteria for MCID. Mean questionnaire end scores are summarized in Supplementary Materials Table 4. The average questionnaire score for the IDS-C at baseline was 31.33 (SD=12.48) and 26.00 (SD=13.34) at visit 2, reflecting a mean score change of -2.55 (SD=14.98, p=0.44) during the study. The average questionnaire score for the QIDS at baseline was 14.60 (SD=5.38) and 10.30 (SD=6.38) at visit two, reflecting a mean score change of -2.63 (SD=4.14, p=0.12) during the study. A Wilcoxon rank sum test was performed examining changes in depression (PHQ-9, IDSC, QIDS-SR-16) and anxiety (GAD-7) from baseline to study exit and the score changes for all questionnaires were found to be statistically insignificant at ɑ=0.05 (see Supplementary Materials Table 4).

In addition, the baseline visit was conducted without the tool, and in general patients did not have significant changes in treatment during Visit 1 or Visit 2 that the AI or CDSS could have influenced. In addition, any changes made at visit 2 would have had their effects after the end of the study. As such it is not possible to comment on the impact of the CDSS on treatment effectiveness; rather, the aim of the study was to evaluate feasibility and not clinical effectiveness. This is reflected in the pre-specified study aims and hypotheses.

LIMITATIONS:

Additionally, COVID-19 led to a mix of telemedicine and in-person appointments. Eight of the 15 non-initial intake baseline appointments were online or over the phone, while the rest were in person. As such, it is not possible to conclude definitively that the results would be the same when looking at only in-person appointments. That being said, the appointment times themselves - lasting roughly 20 minutes - are reasonable lengths in terms of usual clinical practice; in addition, the mean appointment lengths between in-person and virtual appointments at baseline were similar (see Figure 2). As such, this may not be a significant limitation, though future research will be required to confirm this.

| **How much of a threat, if any, is the coronavirus outbreak for:** | **Response Options** | **N=10** | **%** |
| --- | --- | --- | --- |
| Your personal health | Not a threat  Minor threat  Moderate threat  Major threat | 2  4  4  0 | 20%  40%  40%  0% |
| The health of the Canadian population as a whole | Not a threat  Minor threat  Moderate threat  Major threat | 0  2  4  4 | 0%  2%  40%  40% |
| Your personal financial safety | Not a threat  Minor threat  Moderate threat  Major threat | 1  8  1  0 | 10%  80%  10%  0% |
| The Canadian economy | Not a threat  Minor threat  Moderate threat  Major threat | 8  1  1  0 | 80%  10%  10%  0% |
| Day-to-day life in your local community | Not a threat  Minor threat  Moderate threat  Major threat | 0  6  1  3 | 0%  60%  10%  30% |

**Table 1.** Patient COVID-19 Threat Scale. Four patients did not complete the questionnaire.

| **How much of a threat, if any, is the coronavirus outbreak for:** | **Response Options** | **N=5** | **%** |
| --- | --- | --- | --- |
| Your personal health | Not a threat  Minor threat  Moderate threat  Major threat | 0  2  2  1 | 0%  40%  40%  20% |
| The health of the Canadian population as a whole | Not a threat  Minor threat  Moderate threat  Major threat | 0  0  2  3 | 0%  0%  40%  60% |
| Your personal financial safety | Not a threat  Minor threat  Moderate threat  Major threat | 3  1  1  0 | 60%  20%  20%  0% |
| The Canadian economy | Not a threat  Minor threat  Moderate threat  Major threat | 0  0  1  4 | 0%  0%  20%  40% |
| Day-to-day life in your local community | Not a threat  Minor threat  Moderate threat  Major threat | 0  0  2  3 | 0%  0%  40%  60% |

**Table 2.** Physician COVID-19 Threat Scale. Two physicians did not complete the questionnaire.

| Side Effect | Patients who experienced (%) | Tolerable (%) | Distressing (%) |
| --- | --- | --- | --- |
| Dry mouth | 85.71 | 83.33 | 16.67 |
| Nausea | 78.57 | 45.45 | 54.55 |
| Diarrhea | 78.57 | 81.81 | 18.19 |
| Constipation | 78.57 | 72.72 | 27.28 |
| Dizziness | 78.57 | 63.63 | 36.37 |
| Palpitations | 64.28 | 33.33 | 66.67 |
| Sweating | 57.14 | 37.50 | 62.50 |
| Headache | 92.85 | 61.53 | 38.47 |
| Tremors | 50.00 | 57.14 | 42.86 |
| Loss of sexual desire | 78.57 | 45.45 | 54.55 |
| Trouble achieving orgasm | 50.00 | 28.57 | 71.43 |
| Trouble with erections | 14.28 | 50.00 | 50.00 |
| Increased appetite | 42.85 | 33.33 | 66.67 |
| Increased weight | 42.85 | 33.33 | 66.67 |
| Emotional indifference | 72.48 | 40.00 | 60.00 |
| Difficulty sleeping: too little | 92.85 | 46.15 | 53.85 |
| Difficulty sleep: too much | 78.57 | 45.45 | 54.55 |
| Anxiety | 100.00 | 35.71 | 64.29 |
| Restlessness | 92.85 | 46.15 | 53.85 |
| Decreased energy | 92.85 | 30.76 | 69.24 |

**Table 3.** Patient PRISE-20 results. All patients completed the PRISE-20 (Patient Related Inventory of Side Effects) Questionnaire. Percentage of patients who experienced each side effect at one point throughout the study is recorded. Percentage of patients who reported the side effect to be either tolerable or distressing is also recorded.

| Mean PHQ-9 End Score | 11.21 (SD = 6.73) |
| --- | --- |
| Mean GAD-7 End Score | 8.07 (SD = 6.55) |
| Mean IDS-C End Score | 26.00 (SD = 13.34) |
| Mean QIDS End Score | 10.30 (SD = 6.38) |
| Mean number of days between baseline and visit 1 | 40.86 (SD = 29.40) |
| Mean number of days between visit 1 and visit 2 | 51.57 (SD = 62.58) |
| Mean baseline visit length (mins) | 19.29 (SD=5.75) |
| Mean visit 1 length (mins) | 17.80 (SD = 9.74 |
| Mean visit 2 length (mins) | 21.39 (SD = 10.28) |
| Trust of the CDSS - indicated by a 4 or 5 on a 5-Point Likert Scale | Rated by Patients (n=13): 61.54%  Rated by Physicians (n=7): 71.43% |
| Ease of use - indicated by a 4 or 5 on a 5-Point Likert Scale | Rated by Patients (n=13): 92.31%  Rated by Physicians (n=7): 71.43% |

**Table 4.** Table of Questionnaire End Scores and Other Study Results.
